# Impact of COVID-19 pre-test probability on positive predictive value of high cycle threshold SARS-CoV-2 real-time reverse transcription PCR test results

**DOI:** 10.1101/2021.03.02.21252768

**Authors:** Jonathan B. Gubbay, Heather Rilkoff, Heather L. Kristjanson, Jessica D. Forbes, Michelle Murti, AliReza Eshaghi, George Broukhanski, Antoine Corbeil, Nahuel Fittipaldi, Jessica P. Hopkins, Erik Kristjanson, Julianne V. Kus, Liane Macdonald, Anna Majury, Gustavo V Mallo, Tony Mazzulli, Roberto G. Melano, Romy Olsha, Stephen J. Perusini, Vanessa Tran, Vanessa G Allen, Samir N Patel

## Abstract

**Background:** Performance characteristics of SARS-CoV-2 nucleic acid detection assays are understudied within contexts of low pre-test probability, including screening asymptomatic persons without epidemiological links to confirmed cases, or asymptomatic surveillance testing. SARS-CoV-2 detection without symptoms may represent resolved infection with persistent RNA shedding, presymptomatic or asymptomatic infection, or a false positive test. This study assessed clinical specificity of SARS-CoV-2 real-time reverse transcription polymerase chain reaction (rRT-PCR) assays by retesting positive specimens from five pre-test probability groups ranging from high to low with an alternate assay.

**Materials and Methods:** A total of 122 rRT-PCR positive specimens collected from unique patients between March and July 2020 were retested using a laboratory-developed nested RT-PCR assay targeting the RNA-dependent RNA polymerase (RdRp) gene followed by Sanger sequencing.

**Results:** Significantly less positive results in the lowest pre-test probability group (facilities with institution-wide screening having ≤ 3 positive asymptomatic cases) were reproduced with the nested RdRp gene RT-PCR assay than in all other groups combined (5/32, 15·6% vs 61/90, 68%; p <0·0001), and in each subgroup with higher pre-test probability (individual subgroup range 50·0% to 85·0%).

**Conclusions:** A higher proportion of false-positive test results are likely with lower pre-test probability. Positive SARS-CoV-2 PCR results should be interpreted within the context of patient history, clinical setting, known exposure, and estimated community disease prevalence. Large-scale SARS-CoV-2 screening testing initiatives among low pre-test probability populations should be evaluated thoroughly prior to implementation given the risk of false positives and consequent potential for harm at the individual and population level.

## INTRODUCTION

The COVID-19 pandemic has resulted in implementation of large-scale testing practices globally with the aim of early case detection. Prompt testing facilitates timely public health and clinical management, including case identification, contact tracing, and treatment. Widespread laboratory testing has also significantly contributed to knowledge of the epidemiology of the SARS-CoV-2 virus. One such observation is that of positive real-time reverse transcription PCR (rRT-PCR) tests in persons without symptoms. This may be represent resolved infections with persistent viral RNA shedding, active presymptomatic (patients who later develop symptoms) or asymptomatic (patients who never develop symptoms prior to or following testing) infections, or because of false positive laboratory tests.^1^ The likelihood of a false positive rRT-PCR result increases as pre-test probability of the condition it is designed to detect decreases. Examples of low pre-test probability scenarios include asymptomatic groups with no known exposure to COVID-19 cases, and communities with low prevalence of COVID-19. Further, a positive rRT-PCR result close to the limit of detection (LOD) of an assay has a greater likelihood of being false positive.^2^

False positive results can be attributable to pre-analytical errors (e.g. specimen contamination or aliquoting errors), analytical errors (e.g. quality assurance failures, reagent contamination, or non-specific assay signal detection), or post-analytical errors (e.g. improper interpretation or transcription of results). As outlined by Cohen and Kessel, false positive results can have unintended consequences on the public health response including outbreak identification and modelling, case reporting, and resource allocation.^3^ Moreover, negative consequences are greatest when disease prevalence is low. Therefore, pre-test probability is important to consider when discriminating between true or false positive rRT-PCR results.

The cycle threshold (Ct) value, an indirect measure of viral load, and its application to test interpretation has become an important tool for public health practitioners. Together with available clinical and epidemiological factors, the Ct value can help determine appropriate public health follow-up (e.g., contact tracing and/or outbreak declaration) for asymptomatic patients.^4^ However, multiple studies have shown that Ct values overlap between symptomatic, presymptomatic, and asymptomatic cases, and that time from initial infection to testing is the most significant determinant of Ct value.^5-7^ Presymptomatic persons may have comparable viral loads to symptomatic individuals and may be just as likely as symptomatic individuals to infect others due to their unawareness of having the virus, hence their identification has far reaching implications for public health management .^8,9^

Ontario is the most populous province in Canada with approximately 14.7 million residents^10^. The first case of COVID-19 in the province was identified in a patient who presented to hospital on January 23, 2020.^11^ The first pandemic wave peaked in April and was characterized by a disproportionate impact on congregate settings including residents in long-term care and retirement homes and for some workplaces.^12^ During the first wave, policy changes expanded testing to asymptomatic screening programs within long-term care, hospitals, and some workplace settings, irrespective of risk factors or community prevalence. Such widespread testing has brought into focus the interpretation and implications of positive SARS-CoV-2 rRT-PCR results with high Ct values, since many of these settings have both low prevalence and low pre-test probability of infection.

This study evaluates the relative burden of false positive testing outcomes when testing persons in low pre-test probability settings, by exploring the likelihood of a reproducible positive test result upon retesting specimens having high rRT-PCR Ct values, stratified by pre-test probability. The findings have the potential to inform testing guidelines for persons with low pre-test probability, such as within low prevalence communities.

## MATERIALS AND METHODS

Public Health Ontario (PHO) Laboratory, the Ontario provincial public health and reference laboratory, conducts approximately 25% of SARS-CoV-2 testing for the province. Specimens are submitted from acute care (e.g., hospitals), community, institutional, and occupational settings, as well as from outbreaks. Specimen data were obtained from the PHO laboratory information system (LIS).

A total of 122 specimens from unique patients aged 10 to 99 years (median 53.5 years) who underwent clinical testing between mid-March and July 2020 were included in the analysis. All specimens included were initially positive by rRT-PCR with Ct value ≥ 35 using either (i) a laboratory-developed test (LDT) targeting the envelope (E) gene, or (ii) a commercial assay targeting the E and open reading frame 1ab (ORF1ab) genes (cobas® SARS-CoV-2, Roche Diagnostics, Germany).^13^

Interpreting results for specimens tested using the LDT E gene rRT-PCR assay was based on prior validation data, which determined a LOD of 192 copies/ml of primary sample (95% CI 16 to 2,392 copies/ml of specimen), corresponding to Ct values between 34·8 and 38·7. Based on this data, LDT Ct results ≤38·0 are reported as detected and Ct values ≥40·0 are reported as not detected. Ct values between 38·1 and 39·9 are reported as indeterminate.^14^ Indeterminate results may be due to low viral target quantity in the clinical specimen approaching the assay LOD, failed automated viral RNA extraction, or, in rare cases, nonspecific reactivity (false signal) in the specimen. When clinically relevant or important to public health management, repeat testing is recommended.

Specimens tested with the cobas® SARS-CoV-2 rRT-PCR assay were reported as detected or not detected - the manufacturer does not include an indeterminate range. The maximum number of cycles of PCR amplification used in the assay is proprietary and not provided in the kit insert or other available documentation.

To be included in the study, specimens had to be collected from persons belonging to one of five groups based on information provided on the PHO laboratory requisition. The groups had differing pre-test probability of COVID-19 infection (based on presence of symptoms, prior laboratory confirmation as a case, and epidemiological links to other positive cases) and a high Ct value of ≥35 on either the LDT or cobas® rRT-PCR assay (E gene target). A Ct value of ≥35 was chosen as a conservative estimate of lack of infectivity based on other studies using different assays reporting that a Ct of >34 indicates an individual is not likely to be infectious at the time of diagnostic testing.^7,9,15^ Table 1 contains a description of the five categories, ordered from highest pre-test probability setting (Group 1) to lowest pre-test probability setting (Group 5). Groups 1-4 were tested throughout the study period (March - June 2020), whereas Group 5 was tested beyond the peak of the first pandemic wave (May - July 2020).

**Table 1:**
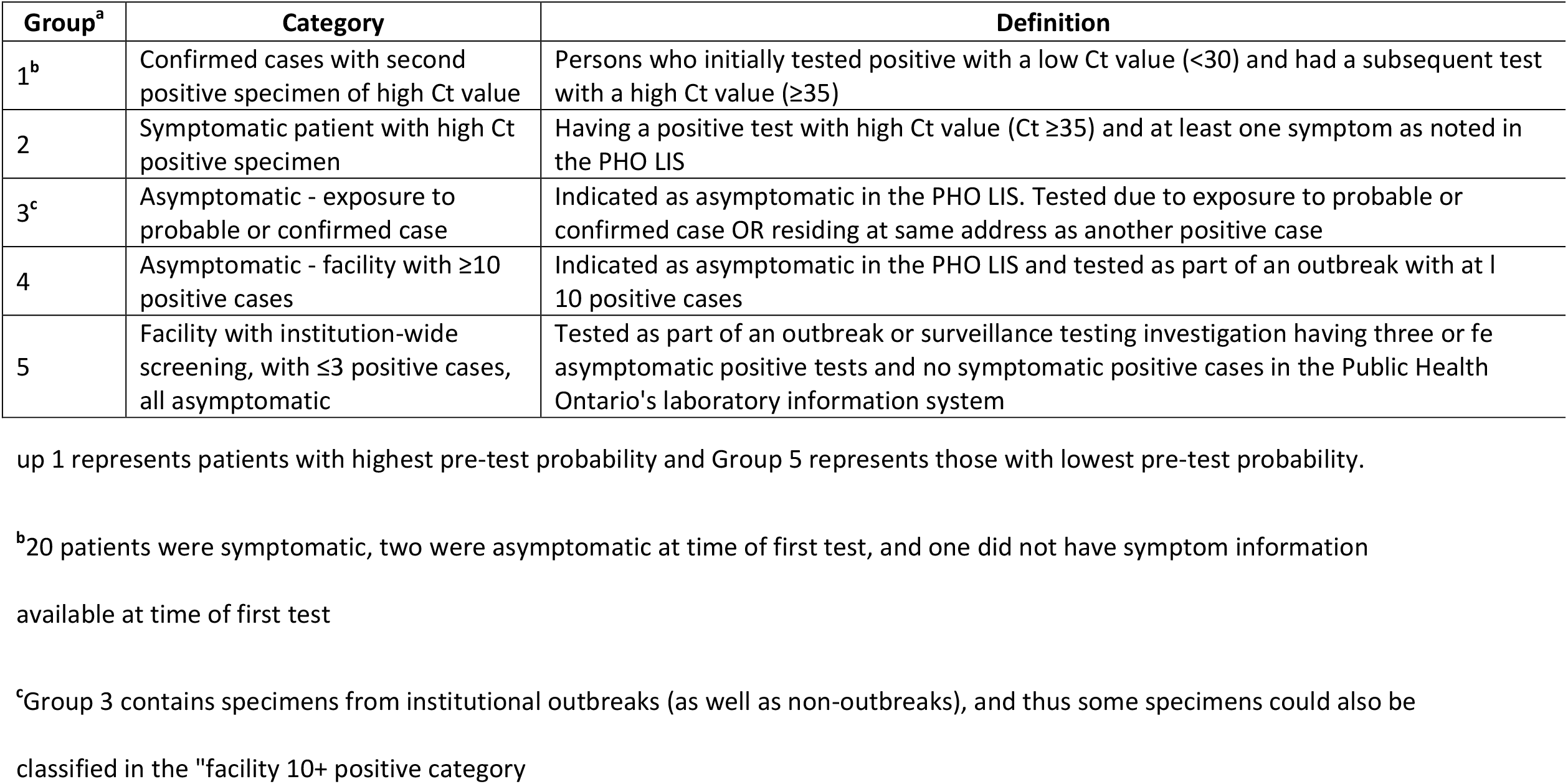
Study Patient Categories and Definitions.

The study dataset was produced by manually reviewing a line list of positive specimens of appropriate Ct values available at the PHO Laboratory, Toronto location, which met inclusion criteria. Group 1, with highest pre-test probability, consisted of high Ct positive specimens from persons with a previous positive result with a lower Ct value (Ct <30). Group 5, which consisted of asymptomatic positives in facilities that underwent whole facility screening with three or fewer positive cases identified, was considered the category with lowest pre-test probability of infection (based on both asymptomatic status and being least likely to be exposed to a case). None of the facilities in Group 5 were in outbreak at the time screening was commenced, which was confirmed by review of the provincial public health information system for the reporting and surveillance of diseases. Group 5 was thus chosen as the reference group when conducting statistical analysis.

Specimens included in this study were retested with a LDT end-point nested RT-PCR assay targeting the RNA dependent RNA polymerase (RdRp) gene, followed by Sanger sequencing of amplicons with expected size of 192 base pairs. This assay was adapted from a previously published Middle East Respiratory Syndrome Coronavirus (MERS-CoV) nested PCR, altered such that the relevant primer bases match SARS-CoV-2: an outer primer and newly designed inner primers were used for both amplification and sequencing.^16^ Primers used are presented in Table 2. The LOD determined during validation was similar to that of the E gene rRT-PCR, at 256 copies/ml of primary specimen (95% CI 37.92 to 1733 copies/ml).

**TABLE 2:**
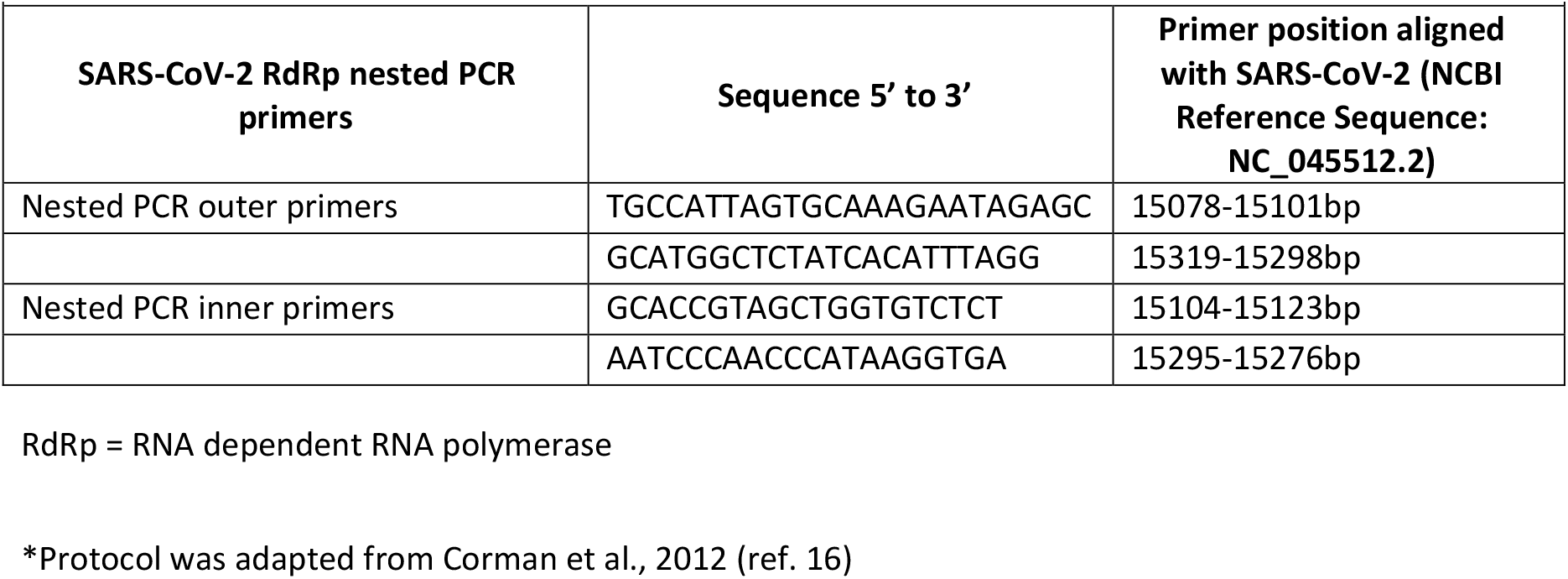
SARS-COV-2 Laboratory Developed RdRp Nested PCR Protocol in Use at PHO Laboratory *.

The proportion of specimens detected by the RdRp gene nested PCR assay as well as the median and range of Ct values were determined overall and for each of the five categories. Statistical significance for median Ct value comparison was calculated using Wilcoxon rank-sum test. Statistical significance for categorical results was calculated using Fisher’s exact test with Bonferroni correction to adjust for multiple comparisons. Results were considered significant at a level of 0·05. All analyses were conducted using SAS Enterprise Guide 8.3.^17^The PHO Ethics Review Board determined that this project is exempt from research ethics committee review, as it describes analyses that were completed at PHO Laboratory as part of routine clinical respiratory testing during the first wave of the COVID-19 pandemic in Ontario and are therefore considered public health practice, not research.

## RESULTS

Table 3 describes the results of the specimens overall and by group. After retesting using the RdRp gene nested PCR assay with Sanger sequencing, results varied according to pre-test probability. Overall, 66/122 (54·1%) specimens had RdRp gene detected. There was a significant difference (p<0·01) in the E gene Ct values among specimens that were reproducible in the RdRp gene nested RT-PCR (median Ct 36·2, range 35·0 to 40·6) compared to those that could not be confirmed (median Ct 37·5, range 35·2-39·8).

**TABLE 3:** Initial E Gene Real-Time PCR and RdRp PCR Results Stratified by Patient Category.

|  |  |  |  |  |  |  |  |  |  |  |  |
| --- | --- | --- | --- | --- | --- | --- | --- | --- | --- | --- | --- |
|  |  |  | PATIENTS |  |  | DETECTED BY RdRp PCR |  |  | NOT DETECTED BY RdRp PCR |  |  |
| Group<sup>a</sup> |  | N | Median Age (Range) | Median Ct (Range) on initial E gene PCR |  | N (%) | Median Ct (Range) on initial E gene PCR |  | N (%) | Median Ct (Range) on initial E gene PCR | P-value<sup>b</sup> |
| 1 |  | 23 | 52 (14-99) | 36.9 (35.0-38.4) |  | 18 (78.3) | 36.7 (35.0-38.3) |  | 5 (21.7) | 38.1 (35.9-38.4) | <0.0001 |
| 2 |  | 20 | 68.5 (26-94) | 36.6 (35.0-38.3) |  | 17 (85.0) | 36.9 (35.03-38.3) |  | 3 (15.0) | 36.3 (35.6-37.4) | <0.0001 |
| 3 |  | 15 | 38 (10-93) | 36.1 (35.4-38.0) |  | 10 (66.7) | 36.0 (35.4-37.2) |  | 5 (33.3) | 37.5 (36.0-38.0) | 0.0078 |
| 4 |  | 32 | 57.5 (15-97) | 37.5 (35.4-40.6) |  | 16 (50.0) | 36.6 (35.4-40.6) |  | 16 (50.0) | 37.7 (35.5-38.3) | 0.035 |
| 5 |  | 32 | 46 (17-95) | 36.9 (35.2-39.8) |  | 5 (15.6) | 36.2 (35.6-37.5) |  | 27 (84.3) | 37.0 (35.2-39.8) | (ref)<sup>c</sup> |
| Total |  | 122 | 53.5 (10-99) | 36.9 (35.0-40.6) |  | 66 (54.1) | 36.2 (35.0-40.6) |  | 56 (45.9) | 37.5 (35.2-39.8) | <0.0001<sup>d</sup> |

<sup>a</sup>Refer to Table 1 for Group definitions
<sup>b</sup>P-values compare proportion detected in each Group to Group 5, as the reference group.
<sup>c</sup>Represents reference group to which other groups are compared
<sup>d</sup>P-value compares Groups 1 to 4 combined to Group 5, as the reference group
Ct = cycle threshold; E = envelope gene real-time Reverse-Transcription PCR; RdRp = RNA dependent RNA polymerase gene endpoint PCR
with Sanger sequencing confirmation.

A similar high proportion of specimens from Groups 1 (18/23; 78·3%) and 2 (17/20; 85·0%), both high pre-test probability groups, had reproducible positive results using the RdRp gene nested PCR assay. However, a significantly lower proportion of positive results in the lowest pre-test probability group (Group 5) were reproducible with RdRp gene nested PCR with Sanger sequencing than in all other groups combined (5/32, 15·6% vs 61/90, 68%; p <0·0001), and in each subgroup.

## DISCUSSION

This study was conducted to ascertain impact of different pre-test probability patient categories on ability to reproduce positive rRT-PCR results of high Ct value (Ct ≥35) with a laboratory developed nested PCR assay targeting an independent gene target, RdRp, including Sanger sequencing of the PCR amplicon. We documented a much lower rate of reproducible positive tests among patients in the lowest pre-test probability category of asymptomatic persons within an institution with three or fewer positive patients identified through screening testing (Group 5). Among this group, only five (15·6%) of 32 patients tested positive with the RdRp gene nested RT-PCR. This is in contrast to the other patient groups with higher pre-test probability, where 50% to 85% of E gene rRT-PCR positive specimens were also RdRp gene PCR positive.

Results of the group-specific analysis suggest that screening asymptomatic individuals in low prevalence situations can generate results which are false positive due to low positive predictive value. This is indicated by the higher percentage of reproducible positive results in symptomatic cases (85·0% detected) compared to cases with no known evidence of disease and likely no contact with positive cases (15·6% detected).

A significantly higher rate of RdRp gene nested RT-PCR positivity was also found among asymptomatic patients exposed to a probable or confirmed case (Group 3, 66·7% detected), and among asymptomatic persons who were tested in facilities with ≥10 positive cases (Group 4, 50% detected), both reflecting higher pre-test probability scenarios than Group 5.

It should be noted that lack of detection with the RdRp gene nested PCR assay is not necessarily a false positive E gene rRT-PCR result, and should not be used to definitively infer false positivity at the individual level. In general, specimens with Ct values well below the assay cut-off for positivity (e.g. Ct values <35 with the positivity cut-off set at Ct 38.0) are less likely to be false positive. However, if the initial positive result was of high Ct value, near the assay cut-off, then repeat testing of the same specimen may yield a negative result, as assay performance near the LOD is not consistent, with results varying on repeat testing. Furthermore, different assays will perform differently on the same specimen having virus quantity near the LOD. However, when applied at a group level, these results provide an indication of the potential relative contribution of false positive test results that may occur in different settings which are differentiated by pre-test probability.

In general, the PPV of COVID-19 PCR assays is excellent among patients with high pre-test probability and approaches 100%.^2^ This was previously determined at PHO using Sanger sequencing of amplicons from RT-PCR positive specimens excluding those for which viral copy number was near the LOD of the assay. However, when testing asymptomatic patients with low pre-test probability in low prevalence settings, the PPV is likely to be different. For example, if community prevalence of SARS-CoV-2 is 1% with a rRT-PCR test sensitivity of 80% and specificity of 99%, the PPV of a positive test is only 44·7%. If prevalence were to increase to 5% or 10%, then the PPV increases significantly to 80·8% and 89·9%, respectively. Recent serosurveys conducted by PHO using residual convenience specimens found a low adjusted monthly seroprevalence of 1·1% among specimens received in June, July and again in August, 2020.^18^ This provides further evidence that Ontario overall was a low prevalence setting for SARS-CoV-2 infection during the study period.

Analysis of results from over 100,000 SARS-CoV-2 tests conducted at PHO Laboratory for asymptomatic screening programs (including long-term care homes, retirement homes, childcare settings, hospitals, settings with migrant workers, and correctional institutions) during the same period as this study identified a positivity rate of 0·2%, (unpublished data). Nearly 70% of positive tests had Ct values ≥35, suggesting that true positivity is likely to be lower, given the potential for false-positive high Ct results in these low-prevalence settings.

Limitations of this study include small sample size and use of a non-randomized sampling method that may limit generalizability of findings. Additionally, all specimens from Groups 2 to 5 included in the study were indicated as the individual’s first positive specimen tested by PHO. However, because PHO Laboratory only conducts a subset of Ontario’s testing (approximately 25%), it is possible that patients had additional positive tests processed at other laboratories, which would increase the pre-test probability of the specimen regardless of the group to which the individual’s sample was assigned. For similar reasons, it is possible that not all positive cases from individual institutions were captured in our study if some testing for additional cases was done at other laboratories or individuals declined testing. To substantiate that the low-prevalence institutional settings had no more than three cases, validation was conducted against provincially reported outbreak-related cases associated with these settings within a three week period. It was assumed that if the number of cases identified by the asymptomatic screening program became greater than 3, an outbreak would be declared and any additional cases would be captured as part of the outbreak.

A further limitation of this study involves changes to government testing guidelines as the pandemic evolved: changes occurred in both the population incidence of COVID-19 as well as guidelines indicating who should be tested during the period of this study, from March to July 2020. This may introduce a bias, since persons tested in periods with more cases, or with stricter testing criteria as occurred earlier in the pandemic in Ontario, may have a greater likelihood of infection.

Specimens included in this study were stored at -80C for weeks to months prior to conducting the RdRp gene nested RT-PCR. RNA degradation during storage and freeze-thaw is possible, and was more likely to affect specimens that were close to the LOD, resulting in a negative RdRp RT-PCR in a specimen that was true rRT-PCR positive at the time of initial testing. Specimen inclusion was based on E gene Ct value at the time of rRT-PCR. Determination of Ct values for LDTs rely on interpretation by the reporting technologist, introducing variability in Ct value assignment. This introduces a risk of reporter bias influencing the specimens included in this study.

Despite these limitations, the results presented here are an important first step towards quantifying the magnitude of false-positive test results in low-prevalence settings, which will increasingly become the norm in many countries with increased use of widespread testing, including broad testing in low pre-test probability populations. Globally, few studies have investigated widespread testing results from low-prevalence areas. This may be attributed to factors such as testing capacity, resources, and practicality. Instead, testing is often restricted to travellers from high prevalence countries, or to those individuals who present with symptoms or a history of exposure, which is linked to public health management.^19^ Currently, there exist a few studies that have attempted to ascertain prevalence through probability-based population-level surveillance studies, rather than initiating a study in an area known to have low prevalence.^20,21^ Examples of targeted low-prevalence studies include examination of potential SARS-CoV-2 detection in wastewater, as well as serosurveillance studies in low prevalence areas.^22,23^

The results of this study have implications for informing future testing approaches, including the utility of conducting broad screening with PCR-based tests in settings with low pre-test probability. For example, in Ontario, this work has been used to inform recent public health approaches, resulting in discontinuation of unnecessary public health management such as case isolation, contact tracing, and outbreak declaration for asymptomatic SARS-CoV-2 rRT-PCR positive persons with low pre-test probability who are negative on retesting.^2^

## CONCLUSIONS

SARS-CoV-2 Ct values can be of use when interpreting positive laboratory results derived from patients with low pre-test probability, in particular asymptomatic persons with no epidemiological link to a confirmed COVID-19 case. Ontario has produced guidance documents facilitating risk-based patient management and follow-up that do not rely on definitively concluding if the specimen is true or false positive, which often cannot be achieved with high Ct-positive specimens due to difficulty in reproducing results near the assay LOD on retesting. Health care providers, public health professionals, policy-makers and the public will benefit from ongoing education to understand that false positive tests will occur when testing asymptomatic patients during periods of low community prevalence of SARS-CoV-2, even when utilizing assays with excellent performance. These false positive tests and unnecessary case isolation, contact tracing and outbreak declaration likely outweigh the benefits from the low numbers of true cases detected among these populations.

## Data Availability

Data requests can be made to the corresponding author for institutional review.

## Notes

## Author contributions

JBG, SNP, and HR conceived the study, with input from MM and JH. AE, SP, GM, and GB conducted laboratory work required for this study. HR verified the underlying data, conducted data collection and data analysis. JBG, HR and HK wrote the initial drafts of the manuscript. All other authors reviewed and edited the manuscript equally.

## Acknowledgements

We gratefully acknowledge the staff of Virus Detection and Molecular Diagnostics, Public Health Ontario Laboratory, for diagnostic testing of SARS-CoV-2 specimens.\

